# Unraveling the impact of COVID-19 on urban mobility: A Causal Machine Learning Analysis of Beijing’s Subway System

**DOI:** 10.1101/2024.08.22.24312324

**Authors:** Linmu Zou, Yanhua Chen, Rui Guo, Peicheng Wang, Yanrong He, Shiyu Chen, Zijia Wang, Jiming Zhu

## Abstract

The COVID-19 pandemic has drastically altered urban travel patterns, particularly in public transportation systems like subways. This study examines the effects of the pandemic on subway ridership in Beijing by analyzing the influence of 19 factors, including demographics, land use, network metrics, and weather conditions, before and during the pandemic. Data was collected from June 2019 and June 2020, covering 335 subway stations and over 258 million trips. Using a three-stage analytical framework—comprising Light Gradient Boosting Machine (LightGBM) for fitting, Meta-Learners for causal analysis, and SHapley Additive exPlanations (SHAP) for interpretation—we observed a substantial decline in ridership, with approximately 10,000 fewer passengers per station daily, especially in densely populated areas. Our findings reveal significant shifts in influential factors such as centrality, housing prices, and restaurant density. The spatiotemporal analysis highlights the dynamic nature of these changes. This study underscores the need for adaptive urban planning and provides insights for public health strategies to enhance urban resilience in future pandemics.

**Significance:** The COVID-19 pandemic has highlighted the vulnerabilities of urban transportation systems, especially subways, to sudden disruptions. This study explores how various factors influencing subway ridership in Beijing changed during the pandemic, revealing significant shifts in travel patterns. By understanding these changes, we can better prepare for future public health emergencies and improve urban resilience. Our research provides critical insights for urban planners, public health officials, and policymakers, enabling them to make informed decisions that enhance the adaptability and sustainability of urban environments in the face of global challenges.

## INTRODUCTION

The outbreak and spread of Coronavirus Disease 2019 (COVID-19) has posed a profound global impact^1,2^ and great challenges to urban public health^3^. Undoubtedly, COVID-19 has changed people’s lifestyles dramatically. In response to the virulent contagion, numerous jurisdictions swiftly enforced strict travel prohibitions and directives for home isolation^4^, leading to dramatic changes in people’s travel behavior^5^. The changes in travel pattern well reflected commuters’ behaviors and preferences in response to the outbreak^6,7^, which, could not only help us understand the immediate impacts of the pandemic but should be employed in future planning and policymaking to improve the resilience and adaptability of urban systems.

The unprecedented nature of the COVID-19 pandemic provides an opportunity to study the effects of unforeseen public health events on public transportation infrastructure and urban travel patterns^8,9^, allowing for a unique understanding of how the pandemic reshapes city travel. However, previous studies investigating the influence of COVID-19 on travel behavior and subway usage only examined a few predictors^10-12^ and often lacked a focused analysis on the changes between pre-pandemic and pandemic periods^13,14^. Therefore, there is an urgent need to identify more significant predictors and their changes in order to help inform evidence-based policies in public health.

Robust research methodologies are essential for estimating the impact of the COVID-19 pandemic on public transport ridership, taking potential confounding effects into account. Approaches such as generalized linear models^11^, geographically weighted regression^15^, and interrupted time-series analysis^12^ had been employed to examine the association between pandemic and commuting trip patterns. However, confounding effects brought by the spatial development of the city and other external factors (e.g. weather conditions) had been overlooked. Little is known about the causal relationship between pandemic and urban travel. Moreover, the precision of previous findings might be attenuated by the stringent asumptions^16-18^, including the unobserved confounding variables, the parallel trends and the stable composition.

To have a comprehensive understanding of how the pandemic and other contributing elements influenced subway travel, we developed an analytical framework to estimate the causal effect of COVID-19 on subway ridership and examined the spatiotemporal heterogeneity in factors that influence ridership patterns. The impact of COVID-19 is multifaceted and comprehensive, involving the occurrence of the pandemic, subsequent increases in infection rates, implementation of government control policies, shifts in social atmosphere, and changes in individual behaviors. The advanced causal inference models and sophisticated machine learning interpretable techniques enabled us to explore the multifaceted impacts of COVID-19.

## METHODS

### Study design and data sources

The Beijing Subway stands as a highly efficient mass transit system catering to the needs of more than 20 million residents. It is one of the most extensive and vibrant urban rail networks globally, covering a vast expanse of more than 720 kilometers and 22 lines by the end of 2020. The spatial distribution of the Beijing rail transit network in 2020 is shown in Supplementary Figure 1. In 2020, 2,294 million passenger journeys were facilitated by the system, marking a 42.10% decline from 2019^19^. The decreased journeys are intricately associated with the outbreak of Xinfadi in Beijing on June 13, 2020. The emergence of a new wave of COVID-19 prompted the Beijing government to implement stringent control policies and define the high-risk regions, which, provided us a good opportunity to study the causal impact of COVID-19 on people’s travel behavior.

Two specific time points were selected to explore the effects of the COVID-19 outbreak on travel behavior: June 2020, the month when the outbreak occurred, and the preceding year without outbreak (June 2019). We collected various data for each station, including the subway ridership, demographics, land use properties, network metrics, and weather conditions. These data allowed for a comprehensive analysis on the factors that influence subway ridership during the pandemic and non-pandemic periods. Supplementary Table 1 shows the characteristics of the overall dataset.

### Outcomes

The main outcome of our study was ridership, measured by the daily traffic volume in stations. Data were collected through the Automatic Fare Collection system (AFC) from Beijing transport authority. We excluded ridership of the airport express line due to its unique passenger transport patterns in the outer city ring, which was different from other local regions in land use and demographics. The final ridership included in this study covered the period before COVID-19 (19 lines with 327 stations, a total of 169,913,012 trips) and during COVID-19 (19 lines with 335 stations, a total of 88,283,273 trips).

### Demographic

The demographic feature of stations was collected from the publicly available spatial demographic dataset WorldPop (https://hub.worldpop.org/), which was created to provide open, high-resolution geospatial population distribution data. Supplementary Figure 2 portrays the population distribution in Beijing for year 2020. We captured population data within an 800-meter radius of each station to represent the demographic characteristics before and during the COVID-19 pandemic, respectively.

### Land use properties

Land use properties are the unique attributes of a specific land area for each station. In this study, we integrated Point of Interest (POI) data, house pricing, and rental value to measure the potential travel demand. We extracted eight POI categories (Bus Stations, Shopping Outlets, Enterprises, Residences, Accommodation Facilities, Hospitals, Restaurants, and Scenic Sites) from the web service API through the AutoNavi open platform (https://developer.amap.com/). House price and rental value were proxies for assessing geographical convenience, surrounding facility amenities, and the intrinsic value of land^20^, which were obtained from the Chinese real estate service platform (https://beijing.anjuke.com/).

### Network metrics

We employed a complex network theory to capture the metrics of each station, which views the lines and stations as edges and nodes within a rail network. Node Importance Indicators (NIIs), such as Betweenness Centrality, Closeness Centrality, Eigenvector Centrality, and PageRank, were used to highlight the pivotal nodes or important stations within the transportation network. A higher NII value indicates increased node importance within the network (Methods Appendix).

### Weather conditions

Weather conditions were obtained from nine meteorological base stations that were close to the Beijing subway managed by the Meteorological Bureau, as shown in Supplementary Figure 1. We assumed that weather conditions at a subway station corresponded to those located at its nearest neighboring meteorological base station. This dataset included four key meteorological parameters: average temperature (°C), average relative humidity (%), average wind speed (m/s), and accumulated precipitation (mm).

### Three-stage analytical framework

To estimate the effect of COVID-19 on city travel, we developed a three-stage framework, including a state-of-the-art machine learning model, the robust data-driven causal inference methods, and the interpretable theories. The theories were used to examine changes in spatial and temporal heterogeneity of factors that influence subway travel ridership before and after the onset of COVID-19 (Figure 1). Technical details are provided in the Methods Appendix.

**Figure 1.**
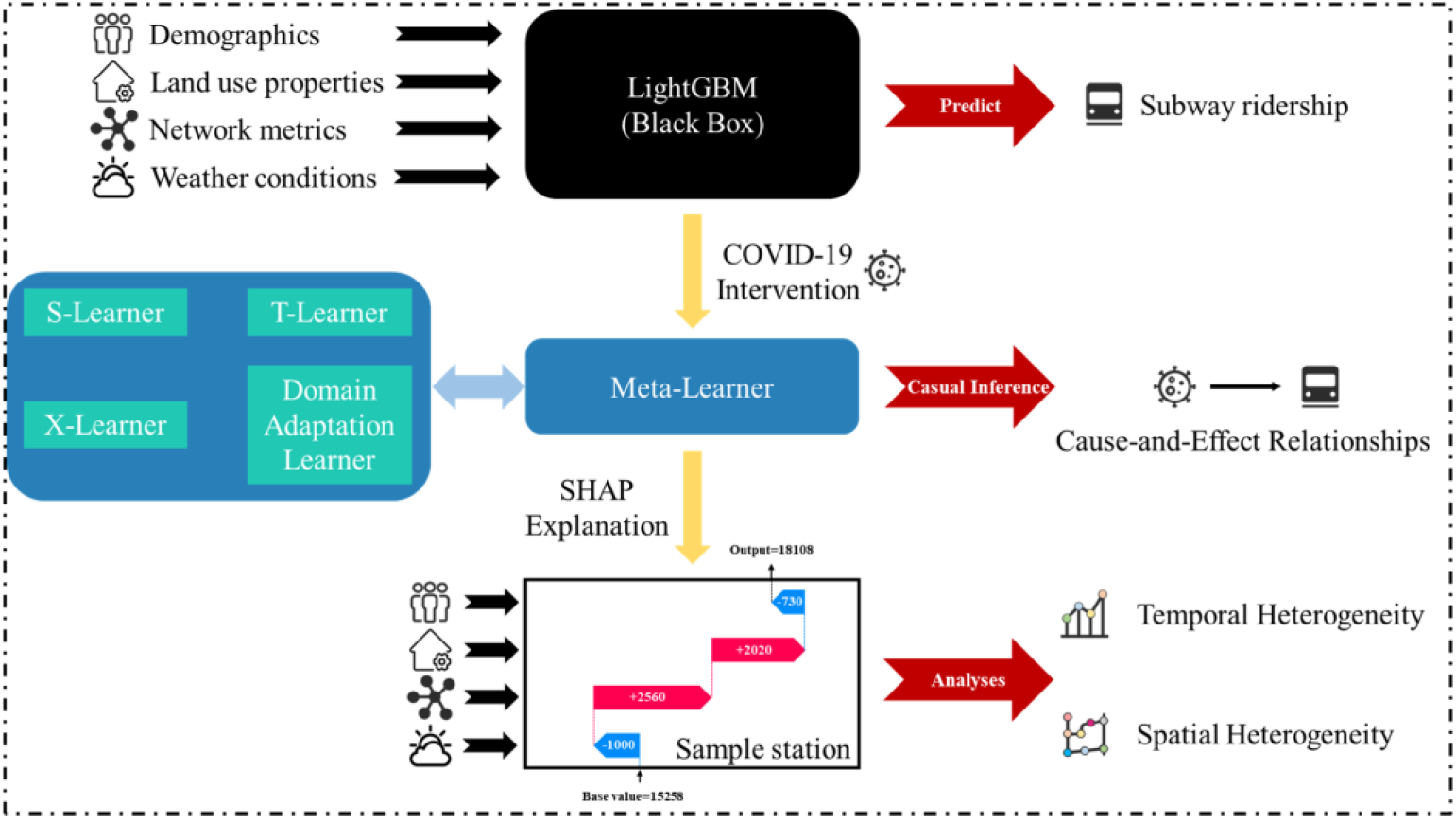
The framework for estimating the effect of COVID-19, covering three key domains: machine learning, causal inference, and interpretable framework. We utilized the LightGBM machine learning algorithm for modeling, employed Meta-Learners to estimate causal inferences, and leveraged SHAP as the interpretable theories.

Stage 1 (machine learning modeling): A Light Gradient Boosting Machine (LightGBM^21^) was introduced to enhance the predictive accuracy of gradient boosting models, particularly in scenarios with high-dimensional input features and substantial data volumes. We employed the LightGBM regressor for regression tasks and the LightGBM classifier for the propensity model, while maintaining a learning rate of 0.1 for both.

Stage 2 (causal effects analysis): Meta-Learners^22^, within the realm of causal inference, were employed as discrete treatment estimators to estimate the Conditional Average Treatment Effect (CATE). These methods address causality in observational data and are crucial in scenarios where randomized controlled trials are limited. Four distinct Meta-Learner methods were utilized: S-Learner, T-Learner, X-Learner, and Domain Adaptation Learner. Our study leveraged LightGBM models for all causal estimations (Models comparison shown in Supplementary Table 2). Subsequently, the concept of permutation importance^23^ was applied to calculate the importance of each variable to the causal estimation.

Stage 3 (complex model interpretation): Despite the computational efficiency and predictive accuracy of machine learning methods, they often function as black-box models that fail to explain the precise impacts of important features on predictions. SHapley Additive exPlanations (SHAP^24^), rooted in game theory, was employed as a post-modeling technique to interpret the outputs of intricate machine learning models. By assigning a value to each input feature, SHAP facilitated an understanding of how and to what degree each feature contributed to the final predicted outcome, providing interpretable explanations for model results (Supplementary Figure 3).

There was no missing data, and all data underwent max-min normalization. The binary cross-entropy loss function was used for the propensity model, estimating the likelihood of a station exposed to the COVID-19 pandemic based on observed characteristics. The root mean squared error loss function was used for the regression tasks. Analyses were conducted using Python V.3.7 and ArcGIS V.10.8.

## RESULTS

### Causal effects of COVID-19 on subway ridership

Compared to the same period in 2019, the number of trips in June 2020 decreased by almost half. As shown in Figure 2, the ridership in Beijing in June 2019 was stable, and fluctuations only occurred in holidays. In December 2019, COVID-19 broke out in China and sporadically in Beijing. The nationwide travel controls resulted in a decrease in ridership compared to the previous year, but still maintained a similar state of equilibrium. However, after the massive outbreak of COVID-19 in Xinfadi, Beijing, the ridership fluctuations became apparent and could be divided into two distinct phases. After 13 June, the ridership volume was unstable and was lower than the average.

**Figure 2.**
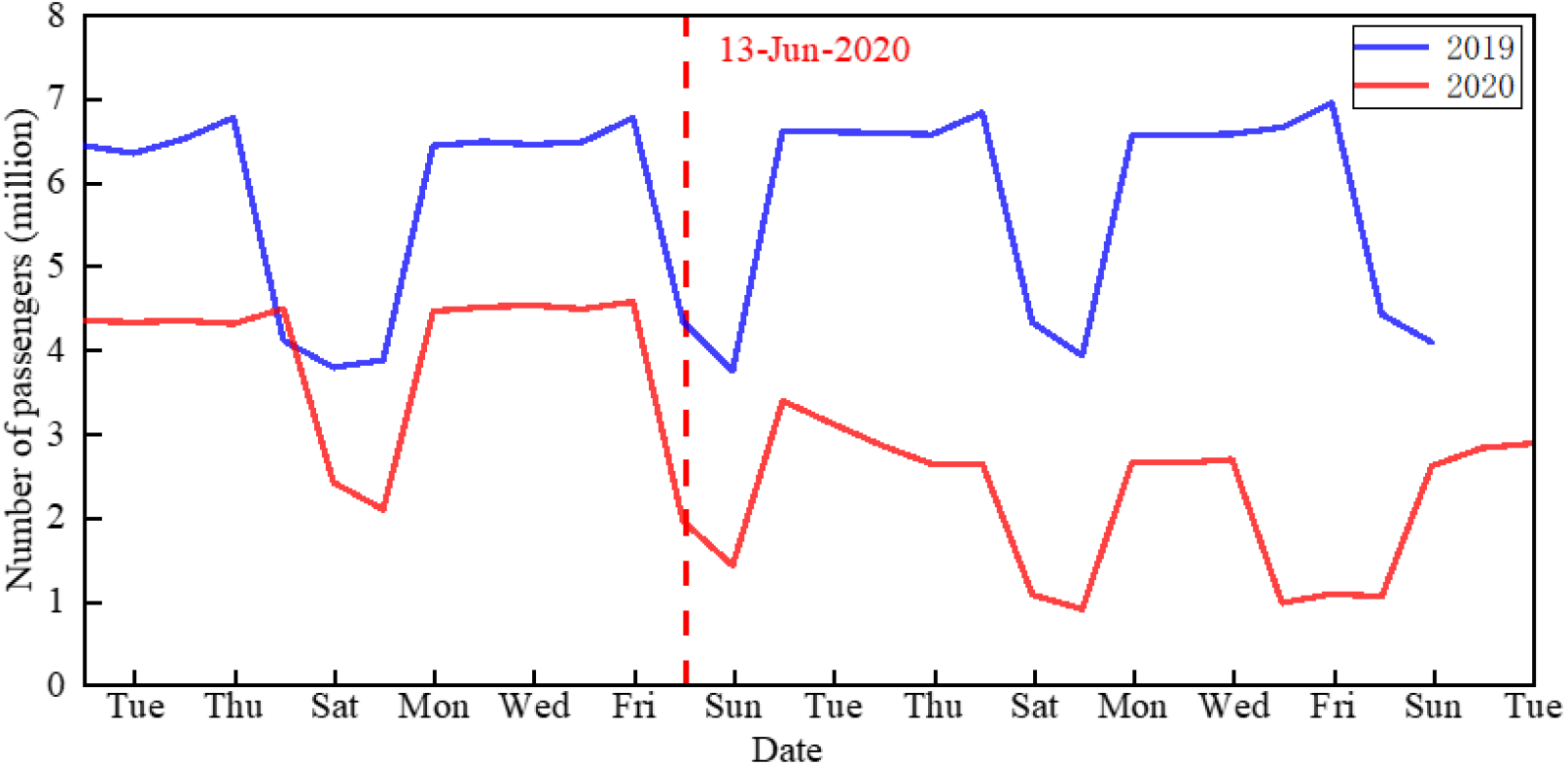
Network-wide ridership for June 2019 and June 2020. The horizontal axis corresponded to the weeks of the two months, and the red vertical line signified the onset of a new epidemic outbreak in June 2020, which was the focal point of our research.

The outbreak in 13 June 2020 was set as the intervening variable to estimate the causal effects of COVID-19 in various meta-learners. Four causal estimators were employed to examine the effect (CATE) of COVID-19 on weekdays (Supplementary Figure 4): S-Learner (ATE = -9484), T-Learner (ATE = -11319), X-Learner (ATE = -10778), and Domain Adaptation Learner (ATE = - 12076). The result showed that the pandemic and related policies led to a decreased tap-in ridership, with approximately 10,000 passengers per day not taking subway during the study period. Stations located near city centers, industrial technology parks, large residential zones, and densely populated suburban regions experienced a more profound impact of COVID-19 than stations in other area. Notably, the result of S-Learner tended to be more conservative than the aggressive one of Domain Adaptation Learner. As a result, we used the X-Learner (Figure 3A) as the focal variable in our analysis.

**Figure 3.**
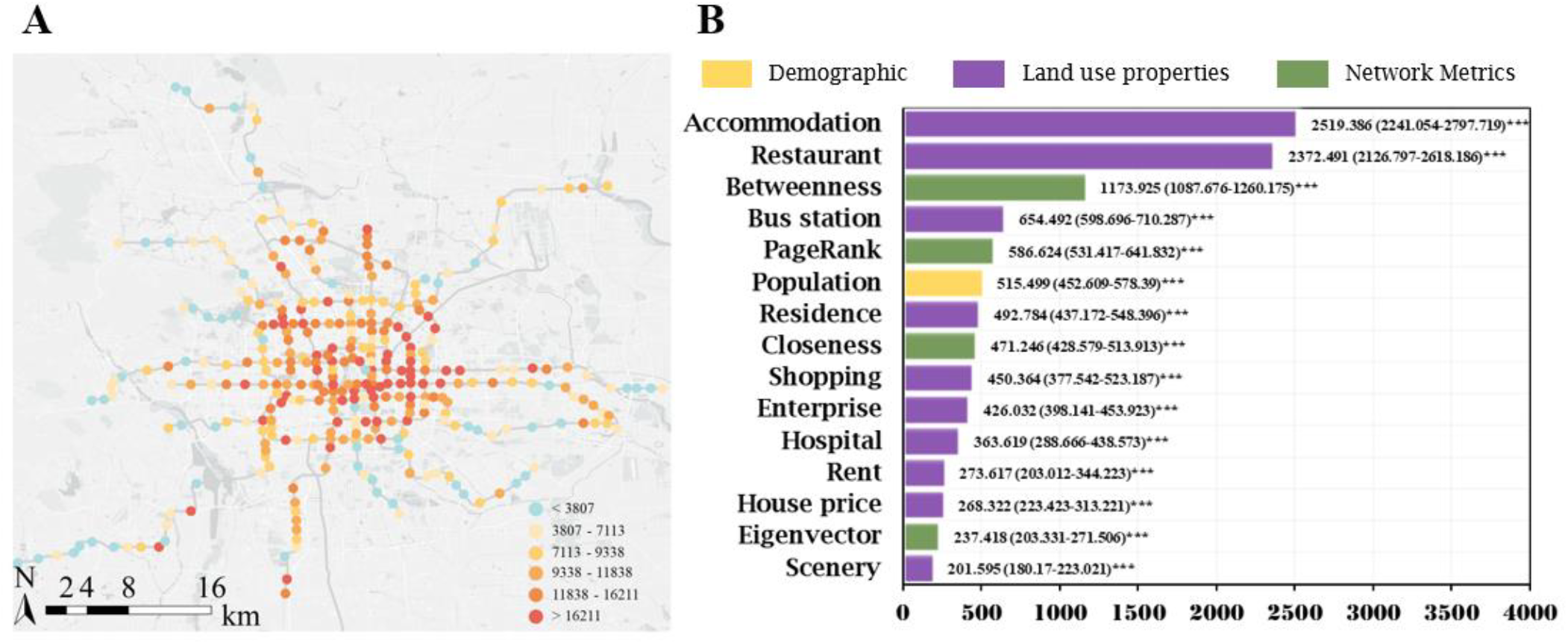
The causal estimates of COVID-19 on subway ridership and their influencing factors. (A) The spatial distribution of causal estimation under X-Learner, indicates the reduction in tap-in ridership as a result of the intervention of COVID-19. (B) Permutation importance scores and Confidence Interval (95% CI). *** means p<0.001 (a p-value of 0.01 indicates that there is a 1% chance that the feature is useless or harmful and a 99% chance that the feature is useful).

The feature importance scores of each feature were then calculated by machine learning regression models (Figure 3B). All features were significantly correlated with the reduced ridership, as all significance values were found to be less than 0.001. The ranking of the feature importance scores showed that the two features with the highest importance scores were accommodation (2519.386, 95%CI 2241.054 to 2797.719) and restaurant (2372.491, 95%CI 2126.797 to 2618.186) and betweenness Centrality (1173.925, 95%CI 1087.676 to 1260.175). In contrast, scenery (201.595, 95%CI 180.170 to 223.021) had the lowest importance score.

### Temporal effects of factors influencing ridership before and during the COVID-19

Based on the SHAP interpretable theory, Figure 4 shows temporal effects of different predictors and their changes before and after the COVID-19 outbreak. Prior to the COVID-19 pandemic, bus station, accommodation, restaurant, enterprise POIs and all four NIIs had a distinctly positive effect on ridership, whereas shopping POIs, scenery POIs, house prices, and rents had a negative impact. During the pandemic, these relationships underwent significant shifts. Further relative importance at two time points for each feature on weekday ridership is shown in Supplementary Table 3.

**Figure 4.**
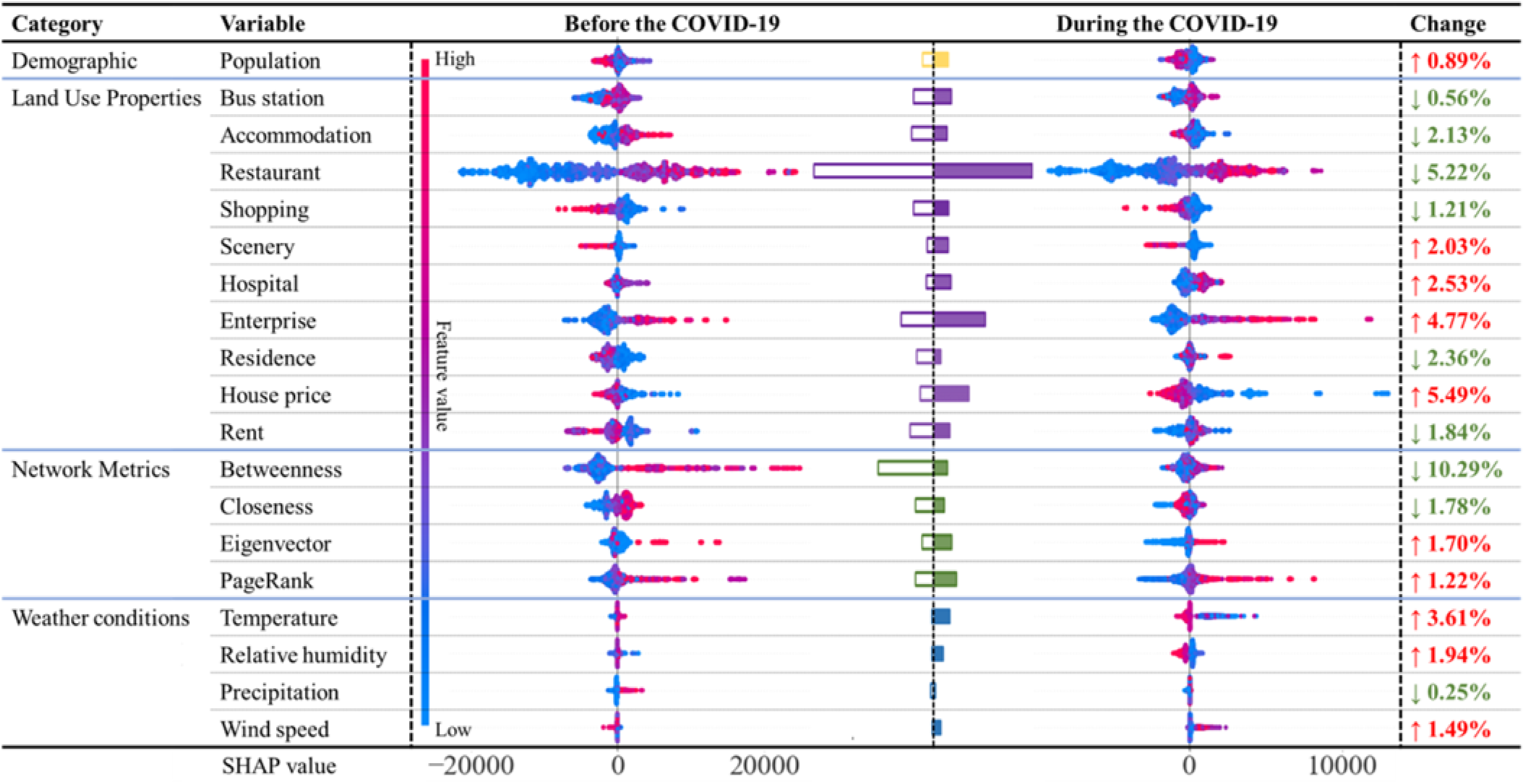
The SHAP summaries plot of all variables and the relative feature importance. The plot conveyed the impact of each sample’s attributes on subway ridership, with the colors denoting the magnitude of the feature value and position reflecting the scale of the SHAP value (Positive and negative SHAP values denote affirmative and adverse contributions to subway ridership, respectively). The histograms visually delineated the shifting relative importance of each variable’s impact on subway ridership, distinguishing between the pre-COVID-19 and COVID-19 periods. The “Change” indicated how much the relative importance had changed before and during the COVID-19.

To be specific, land use prosperities, accommodation (−2.13%), and restaurant (−5.22%) POIs experienced a decline in their positive influence, while enterprise POIs demonstrated an increase (+4.77%). Concurrently, negative relationships also changed, particularly for shopping POIs (−1.21%), scenery POIs (+2.03%), house prices (+5.49%), and rents (−1.84%). It is worth noting that the influence of hospital POIs (1.78%) remained relatively insignificant until the outbreak of COVID-19, whereas during the epidemic this feature had a positive effect and significantly increased +2.53%.

In terms of network metrics, all four NIIs positively contributed to ridership before COVID-19. However, the strong positive impact of betweenness centrality sharply declined during the COVID-19 period (−10.29%). In contrast, closeness (−1.78%), eigenvector (+1.70%), and PageRank (+1.22%) remained relatively stable.

For weather conditions, the importance of temperature (0.45%), relative humidity (0.44%), precipitation (0.67%), and wind speed (0.28%) was less pronounced before the COVID-19 outbreak. However, after the outbreak, the importance of temperature (+3.61%), relative humidity (+1.94%), and wind speed (+1.49%) increased, except for the precipitation (−0.25%) which remained largely unchanged.

### Spatial effects of factors influencing ridership before and during the COVID-19

For spatial variation, we visualized changes in the four most significant features (i.e., betweenness centrality, house price, restaurant, and enterprise), which had the largest magnitude changes before and during the COVID-19 pandemic (Figure 5). Stations where betweenness centrality significantly influenced subway ridership were primarily centralized in the city center before COVID-19. However, during the COVID-19 period, the spatial distribution of these stations became more dispersed. Simultaneously, the impact of house prices on subway ridership gradually diminished in the city center but increased in the suburbs. Similarly, the influence of restaurant POIs on ridership was less pronounced in urban centers, but became more stable in peripheral areas during COVID-19. In contrast, areas influenced by enterprise POIs became more centralized, particularly in key areas such as the central business district (CBD), technology industrial parks, and financial streets.

**Figure 5.**
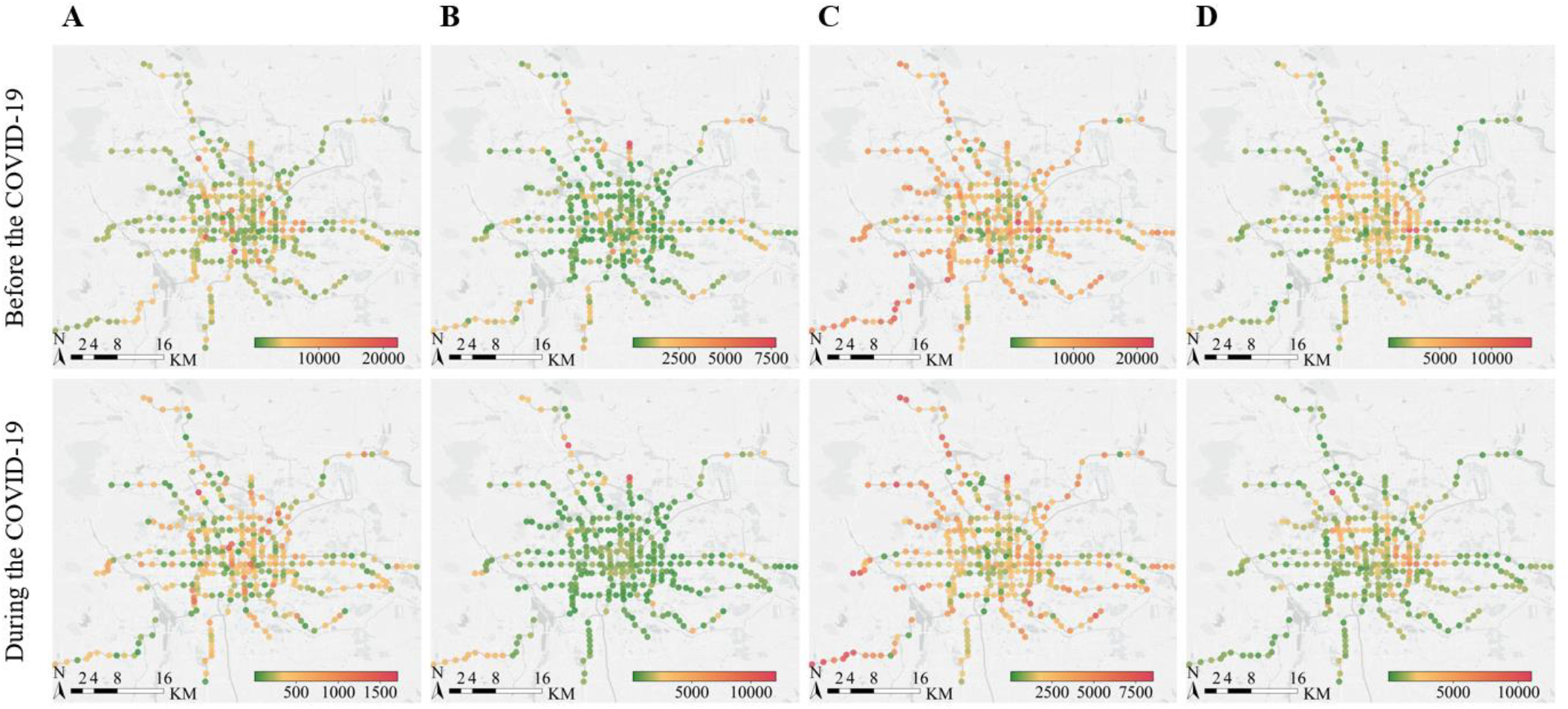
The |SHAP values| for the four variables with the greatest degree of change in relative importance. (A) Betweenness centrality. (B) House price. (C) Restaurant. (D) Enterprise.

## DISCUSSION

Using a three-stage analytical framework based on machine learning approaches and the travel data of subway transport system in Beijing, China, we found a significant reduction in subway ridership (approximately 10,000 passengers per day at each station) due to COVID-19. The comprehensive analysis of various factors yields rich information that should be considered in response to future epidemics.

The impact of COVID-19 was found to be strongest in densely populated areas, which would affect public transportation use. This impact suggests a shift in public travel behavior, steering away from traditional commuting patterns. In response to this change, the transport system needs to redesign the services to align with current demand patterns, as highlighted in previous studies^25,26^. Providing real-time information on crowd density can facilitate an efficient distribution of travel demands, contributing to a safe and responsive transportation system^27^. Meanwhile, the implementation of financial incentives for socially distant transportation alternatives is crucial to compensate for the reduced availability of public transport^26^.

Accommodation and restaurants stand out as the most influential factors in predicting the declined subway ridership, but their positive impacts reduced significantly during the COVID-19 pandemic. This may be resulted from the restrictions on tourism and business travel which led to a reduction of accommodation services^28^. Social distancing measures also affected dining establishments^29^. To address these challenges, outdoor dining and alternative services such as takeout and delivery are recommended^30^. It is also essential to implement enhanced safety measures, including the improved hygiene^31^, a transparent information disclosure for orderly accommodation, and the use of contactless check-in transaction technology. Spatially, restaurant POIs’ importance in central urban area experienced a greater decline compared to the peripheral area. This might be explained by the concentration of high-capacity dining establishments in central area, which leads to the gatherings. In non-central urban areas, the restaurant landscape was more diffuse and resilient, with a slower decrease in the restaurant market^32^. Accordingly, it would be necessary to collaborate with urban planners on zoning scheme plans to reach a balanced distribution of catering venues. This could help mitigate the chain reaction of pathogens and promote a more resilient and resistant urban layout.

During the epidemic, the negative contribution of scenic POIs to tap-in ridership increased, indicating that people may have become more cautious about scenic travel, as these locations are often gathering places but unnecessary destinations. The result aligned with the previous finding^33^ that people’s risk perception of infection significantly influences their willingness to travel. To restore the functionality of scenic areas, it is essential to enhance control over pedestrian flow within each location, ensuring that tourist density remains within safe limits^34^. Additionally, meticulous attention should be given to environmental disinfection and protection measures^31^, especially in indoor attractions.

Our study also observed a positive role played by hospitals who had an increased importance during the COVID-19. The result confirmed previous findings that there was an increased demand for healthcare services and an uptick in hospital visits^35,36^. Consequently, ridership in subway stations near these facilities increased. experienced an increase in ridership. In light of these observations, traffic managers should prioritize traffic management around hospitals^37^. It becomes crucial for hospitals to guide patient flow in an orderly manner, preventing disorder and overcrowding to achieve rational triage during these challenging times. The positive contribution of enterprise POIs to subway ridership also significantly increased and became more concentrated in spatial clusters. This is contrary to the previous finding that stations located in the employment centers were hit the hardest in ridership reduction^38^. The inconsistency of results may be attributed to our more comprehensive indicators measuring land use properties and the application of advanced machine learning inference methods to accurately separate each feature’s nonlinear effects. In fact, considering the reduction of unnecessary trips and the continued necessity of essential commuting^39^, areas with numerous enterprise POIs remained attractive during the COVID-19 period, as suggested by another research^40^. Therefore, to further reduce commute trips, policymakers could consider the promotion of remote work ^41,42^. For work that has to be done offline, flexible work arrangements such as travelling in off-peak hours could be implemented to discourage unnecessary travel and alleviate congestion during peak hours^43^.

We found that the negative association between housing prices and ridership had been strengthened. In general, house prices indicate the socioeconomic status of a neighborhood. The higher price of the land, the greater potential for the presence of high-income groups. The explanation for our finding is that residents in affluent areas typically have a broader range of transportation choices^44^ and were more likely to reduce their reliance on public transport during the pandemic^45,46^. In contrast, people who lived in low-income areas may find it difficult to avoid the use of low-cost commuting options, such as the public transportation^47^. Additionally, our results show that the importance of house prices in the suburbs has intensified more than in the city center. This may be due to the relatively lower incomes and limited transportation options for suburban households. Our findings highlight the divergent travel behavior towards public transportation among different income groups, confirming the heightened vulnerability of low-income population during an epidemic. It is therefore of great necessity to provide alternative transportation options for the vulnerable individuals, enabling them to have access to safe and affordable commuting options^27,48^.

Our results also reveal that the negative contribution of rents to ridership has weakened. This might be resulted from the dynamic nature of rental prices, which act as an indicator of supply and demand equilibrium in the rental market. The weakened relationship reflected a decrease in the transient population^49^ and an accompanying decline in subway ridership, especially during the pandemic when non-Beijing residents were restricted to return^28^. Our study highlights the complexities of immigration during pandemics and calls for the establishment of a more robust real-time monitoring system for the mobile population at a particular time.

In the present study, the biggest drop of importance can be found in betweenness centrality during the epidemic. The feature measures the contribution of each station node to the multitude of shortest paths within the subway network. Stations with high betweenness centrality serve as critical hubs to attract high volumes of passengers. These densely traversed nodes are also more likely to transmit the COVID-19, leading to a higher risk of infection for passengers^12^. The result is consistent with the finding in Shanghai^50^, which found that stations in downtown with high centrality present the highest risk of infection. The impact is also evident in the spatial distribution of passenger congregations; the importance of such stations as collection and dissemination centers was significantly reduced during COVID-19. These results called for a dynamic monitoring of changes in stations, considering their varied centrality during the epidemic and the new cases emerged in the vicinity^51^.

Weather conditions also played a crucial role during the COVID-19 period^52,53^. We found that the impact of weather conditions on subway ridership has increased. This may indicate that passengers have become more sensitive to meteorological situations and preferred to stay indoors during unfavorable weather conditions. The result aligned with previous research^54^ that non-essential trips were more sensitive to weather conditions during the COVID-19 period. Given the weather-induced congregation indoors, our study highlights the importance of improving indoor environmental management to prevent disease transmission^55^, particularly during the pandemic. The provision of real-time and accurate weather information for the public is also essential for them to make appropriate travel choices.

There were some limitations in this study. Firstly, due to the limited acquisition of ridership data from the Beijing transport authority, the study only focused on a narrow timeframe (June 2019 and June 2020) to examine the changes in travel behavior caused by the COVID-19 outbreak. A longer period of data is recommended. Secondly, the study examines a multifaceted and comprehensive impact and therefore does not include dynamic factors such as changing regulations and new virus strains, making it difficult to separate the impact of outbreaks from government interventions. Finally, due to the constraints of data availability, the study only examined population factor as demographic impacts. A more in-depth investigation of societal and economic factors is recommended for future research.

## CONCLUSION

Our study underscores the impact of COVID-19 on Beijing’s transport system. There was a significant decline in subway ridership during the pandemic. The findings indicate that the COVID-19-related interventions have reshaped the city travel patterns, leading to the spatiotemporal heterogeneous variations in factors affecting ridership. Accordingly, collaboration between public health experts, transportation managers, and urban planners becomes imperative to ensure city functioning and the adaption of prevention policies to local conditions. Our study provides valuable insights into pandemic response and helps inform evidence-based strategies to improve urban resilience for the next pandemic.

## Data Availability

All data is included in the article and/or supporting information.

## Data availability

The machine learning models utilized in this paper are publicly accessible through the scikit-learn GitHub repository (https://github.com/scikit-learn/scikit-learn). The causal inference models can be found in the econml GitHub repository (https://github.com/py-why/econml). Additionally, the interpretable techniques are available in the SHAP GitHub repository (https://github.com/shap/shap). All data is included in the article and/or supporting information. All analyses were conducted using the Python V.3.7 and ArcGIS V.10.8.

## Acknowledgements

This research was supported by the National Natural Science Foundation of China (No. 72091514), the Sanming Project of Medicine in Shenzhen Municipality (No. 20212001132), the Bill & Melinda Gates Foundation (No. INV-018302), and the Natural Science Foundation of Beijing Municipality-Fengtai Rail Transit Frontier Research Joint Fund (No. L211002).

