## Supplementary Information for "Unraveling the impact of COVID-19 on urban mobility: A Causal Machine Learning Analysis of Beijing’s Subway System"

### APPENDIX

#### Node Importance Indicators

Betweenness Centrality quantifies a node's importance by enumerating the number of shortest paths that traverse through that specific node. The formula for calculating Betweenness Centrality is as follows:

$$C_b(v) = \sum_{s \neq v \neq t} \frac{\sigma(s,t \mid v)}{\sigma(s,t)} \tag{1}$$

where $\sigma(s,t)$ is the total number of shortest paths between nodes $s$ and $t$, and $\sigma(s,t \mid v)$ represents the number of shortest paths between $s$ and $t$ that pass through node $v$.

Closeness Centrality, gauges how expeditiously a node can reach other nodes within the network. It is computed as the reciprocal of the average shortest path length from a given node to all other nodes. The formula for Closeness Centrality is as follows:

$$C_c(v) = \frac{N-1}{\sum_{s=1}^{N} d(v,s)} \tag{2}$$

where $N$ is the number of stations and $d(v,s)$ represents the shortest path length between node $v$ and $s$.

Eigenvector Centrality, on the other hand, evaluates a node's influence within a network by considering the centrality of its neighboring nodes. It assigns a centrality score to each node in proportion to the sum of the centrality scores of its adjacent nodes. The formula for calculating Eigenvector Centrality is as follows:

$$C_e(v) = \frac{1}{\lambda} \times \sum_{s=1}^{N} A(v,s) \times C_e(s) \tag{3}$$

where $\lambda$ is the largest eigenvalue of the adjacency matrix of the network and $A(v,s)$ denotes the element of the adjacency matrix representing the connection between nodes $v$ and $s$.

PageRank measures the importance or relevance of a web page based on the structure of the web and the links pointing to that page. The underlying idea is that a page is considered more important if it is linked to by other important pages[1]. The formula for PageRank in urban rail network can be summarized as follows:

$$PR(v) = (1-\delta) + \delta \sum_{s=1}^{I(v)} \left( \frac{PR(s)}{I(s)} \times \frac{1}{d(v,s)} \right) \tag{4}$$

where $\delta$ is a damping factor, typically set to around 0.85, and $I(v)$ is the number of outbound links of node $v$. In addition, we use $1/d(v,s)$ as the weight of the link to emphasize the geospatial characteristics of the stations.

#### Light gradient boosting machine

The operational foundation of LightGBM aligns with the fundamentals of gradient boosting[2], and its methodology can be succinctly outlined as follows:

**Leaf-wise Tree Growth:** It presents a distinctive approach to tree construction, distinguishing it from conventional gradient boosting frameworks. Instead of expanding all nodes at a given depth, LightGBM meticulously selects the leaf node that proffers the most substantial reduction in the loss function. This strategy yields the formation of slender and more profound trees, thereby considerably expediting the training process. Moreover, it engenders an ameliorated generalization by yielding shallower trees with diminished node count, thus mitigating the risk of overfitting[3]. Notwithstanding its emphasis on fewer nodes, LightGBM frequently attains a level of accuracy that is competitive or superior in comparison to the traditional depth-wise growth approach.

**Histogram-Based Learning:** This technique discretizes continuous features into discrete bins, thereby transforming the dataset into histograms. Through this transformation, it curtails the computational intricacies associated with gradient computations and facilitates concurrent processing. This strategy engenders a noteworthy expeditiousness in the training process, particularly when dealing with voluminous datasets, thereby contributing significantly to its scalability. Furthermore, it manages memory resources by efficaciously preserving and manipulating these histograms, rendering it eminently suitable for high-dimensional data. The pioneering efforts in the domain of continuous feature handling via histogram-based learning distinctly set it apart as an efficient and scalable gradient boosting framework, ideally tailored for extensive machine learning undertakings.

**Gradient-based One-Side Sampling (GOSS):** It operates through the discerning selection and preservation of instances for training, predicated upon the magnitude of their gradients. It bestows priority upon instances manifesting larger gradients, whilst judiciously subsampling those characterized by more modest gradients. A user-defined ratio confers the ability to regulate the proportion of instances to retain, offering a pliable framework for negotiating the equilibrium between training expeditiousness and model fidelity. GOSS efficaciously ameliorates the proclivity towards overfitting by centering its attention on demanding examples, thus concurrently expediting the training process by dealing with a diminished subset of data points. This technique has verifiably proven its worth in contexts characterized by extensive datasets, thus rendering LightGBM a multifaceted and proficient choice across an array of machine learning tasks.

**Exclusive Feature Bundling (EFB):** It discerns clusters of mutually exclusive features that manifest robust correlations, warranting that only a solitary representative feature from each assemblage garners consideration during tree construction. This curation markedly diminishes the computational overhead associated with the evaluation of prospective splits, culminating in expeditious training intervals and diminished memory requisites. The implementation of EFB assures the selection of the most informative features from each bundle, thereby elevating predictive performance while efficiently managing resources. This feature engineering strategy distinguishes LightGBM as the preeminent choice for tasks entailing datasets replete with an extensive panoply of features.

#### Meta-Learners

**S-Learner:** Employing a "single" estimator, it incorporates the treatment indicator as a feature, akin to the other covariates, without conferring upon it any unique status. The estimated CATE is ascertained by the following expression:

$$\begin{aligned}\hat{\tau}(x) &= E[Y \mid X = x, T = 1] - E[Y \mid X = x, T = 0] \\ &= \hat{\mu}(x,1) - \hat{\mu}(x,0)\end{aligned} \tag{5}$$

where $\hat{\mu} = M(Y \sim (X,T))$ is the outcome model for features $X$, $T$. In this context, $M$ represents any suitable machine learning algorithm.

**T-Learner:** It operates by incorporating "two" functions, namely the control response function and the treatment response function. The estimated CATE is determined as follows:

$$\begin{aligned}\hat{\tau}(x) &= E[Y(1) - Y(0) \mid X = x] \\ &= E[Y(1) \mid X = x] - E[Y(0) \mid X = x] \\ &= \hat{\mu}_1(x) - \hat{\mu}_0(x)\end{aligned} \tag{6}$$

where $\hat{\mu}_0 = M_0(Y^0 \sim X^0)$ and $\hat{\mu}_1 = M_1(Y^1 \sim X^1)$ denote the outcome models for the control and treatment group, respectively. Here, $M_0$ and $M_1$ can be indicative of any suitable machine learning algorithms that possess the capacity to discern and capture the relationship between features and outcomes.

**X-Learner:** It formulates distinct models for $Y(1)$ and $Y(0)$ individually, with the objective of estimating the Conditional Average Treatment Effect on the Treated (CATT) and the Conditional Average Treatment Effect on the Controls (CATC). The CATE estimation for a new data point is derived through a propensity-weighted average of CATT and CATC. A schematic outline of the X-Learner procedure is outlined below:

$$\begin{aligned}\hat{\mu}_0 &= M_1(Y^0 \sim X^0) \\ \hat{\mu}_1 &= M_2(Y^1 \sim X^1) \\ \hat{D}^1 &= Y^1 - \hat{\mu}_0(X^1) \\ \hat{D}^0 &= \hat{\mu}_1(X^0) - Y^0 \\ \hat{\tau}_0 &= M_3(\hat{D}^0 \sim X^0) \\ \hat{\tau}_1 &= M_4(\hat{D}^1 \sim X^1) \\ \hat{\tau} &= g(x)\hat{\tau}_0(x) + (1 - g(x))\hat{\tau}_1(x)\end{aligned} \tag{7}$$

where $g(x) \in [0,1]$ is a weight function, which is an estimation of $P[T = 1 \mid X]$ and $M_1$, $M_2$, $M_3$, $M_4$ stand for the corresponding appropriate machine learning algorithms.

**Domain Adaptation Learner:** It represents a modification of the X-Learner incorporating domain adaptation techniques for the estimation of the outcome models $\hat{\mu}_0$ and $\hat{\mu}_1$. The foundational premise of the Domain Adaptation methodology rests on the assertion that the probability distributions $P(X^0)$ and $P(X^1)$ exhibit disparities[4]. This necessitates the weighting of the $X^0$ samples based on their similarity to the $X^1$ samples during the training of a model on $X^0$ that is unbiased on $X^1$. An outline of the Domain Adaptation Learner procedure is presented below:

$$\begin{aligned}\hat{\mu}_0 &= M_1\left(Y^0 \sim X^0, \text{weights} = \frac{g(X^0)}{1 - g(X^0)}\right) \\ \hat{\mu}_1 &= M_2\left(Y^1 \sim X^1, \text{weights} = \frac{g(X^1)}{1 - g(X^1)}\right) \\ \hat{D}^1 &= Y^1 - \hat{\mu}_0(X^1) \\ \hat{D}^0 &= \hat{\mu}_1(X^0) - Y^0 \\ \hat{\tau} &= M_3(\mathrm{C}(\hat{D}^0, \hat{D}^1) \sim \mathrm{C}(X^0, X^1))\end{aligned} \tag{8}$$

where $g(x) \in [0,1]$ is an estimation of $P[T = 1 \mid X]$ and $M_1$, $M_2$, $M_3$ are suitable machine learning algorithms, and C denotes dataset concatenation.

#### Shapley additive explanations

Let $f(x)$ be a prediction of the original model to be explained, and the explanation model with interpretable inputs $x'$ is expressed as[5]:

$$h(x') = f(x) = \phi_0 + \sum_{i=1}^{K} \phi_i x'_i \tag{9}$$

where $K$ is the number of input features, $\phi_0$ denotes the model output when all interpretable inputs are missing and $\phi_i \in \mathbb{R}$ represents the feature attribution for a feature $i$, also known as the Shapley Value. Combined with the deduction of Shapley[6], the Shapley Value can be calculated as:

$$\phi_i(f, x) = \sum_{z' \subseteq x'} \frac{|z'|!(K - |z'| - 1)!}{K!} [f_x(z') - f_x(z' \setminus i)] \tag{10}$$

where $|z'|$ is the number of non-zero entries in $z'$, and $z' \subseteq x'$ represents all $z'$ vectors where the non-zero entries are a subset of the non-zero entries in $x'$. For a setting $z' \setminus i$ when $z'_i = 0$, there is a solution where $f_x(z') = E[f(z) \mid z_S]$ and $S$ is the set of non-zero indexes in $z'$.

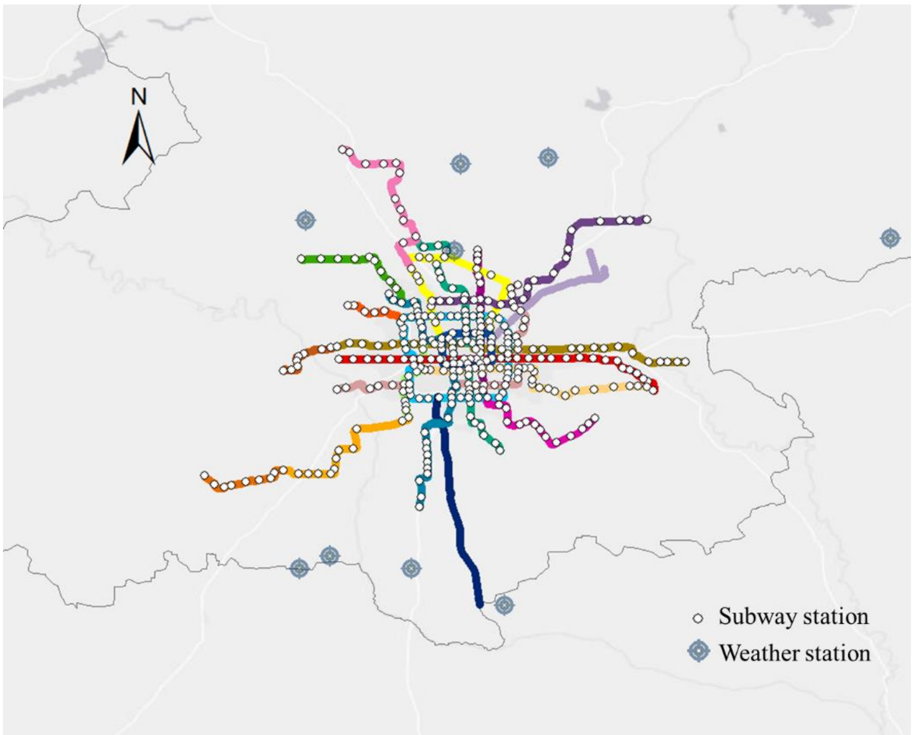

**Supplementary Figure 1:** Distribution of Beijing rail transit and meteorological stations in 2020. Distribution of Beijing rail transit and meteorological stations in 2020. The stations on the airport express line were excluded.

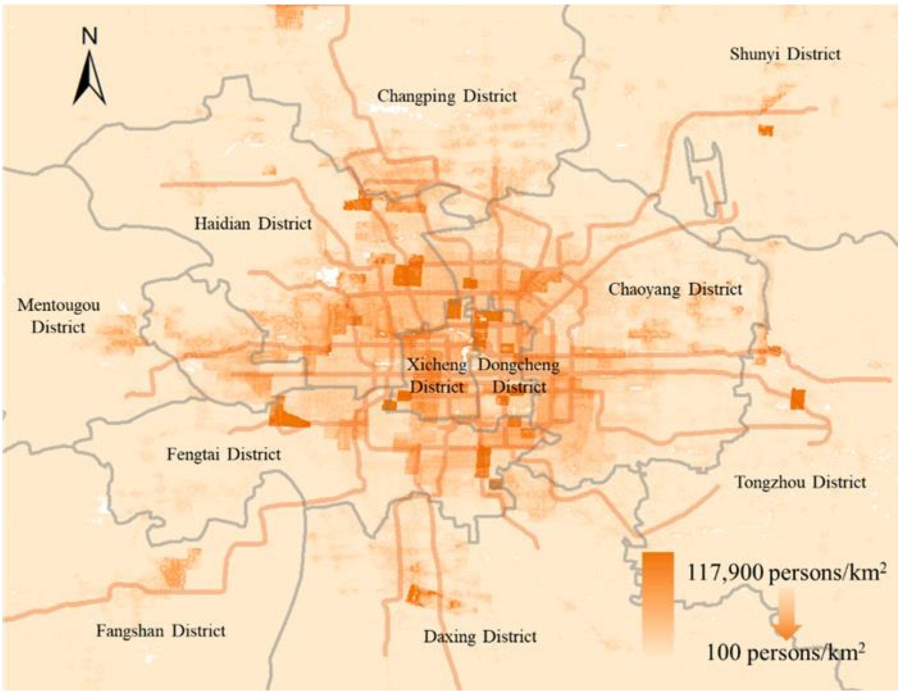

**Supplementary Figure 2:** Population distribution of Beijing in 2020. Darker hues indicate elevated population density.

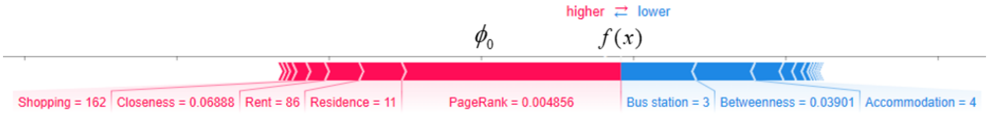

**Supplementary Figure 3:** SHAP values to explain the predicted value of a sample. SHAP values are represented visually as “forces”, where the length of an arrow corresponds to the magnitude of the SHAP value, and these values convey the impact of features. For example, the explained variable (Ridership) commences from the base value $\phi_0$ , and it experiences positive influence from the features depicted in red and negative influence from the features indicated by the blue arrows. In the illustrated sample, it is noteworthy that the presence of three nearby bus stations is deemed to exert a detrimental impact on ridership.

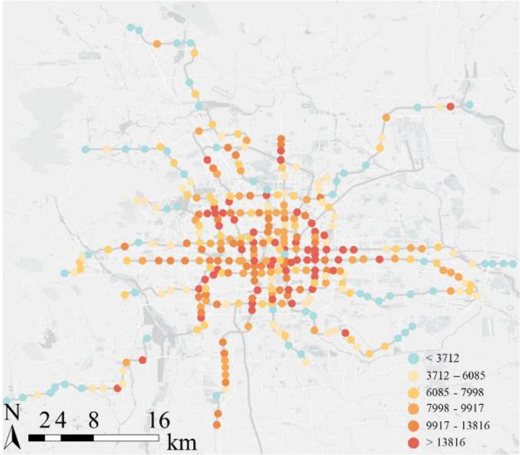

(a)

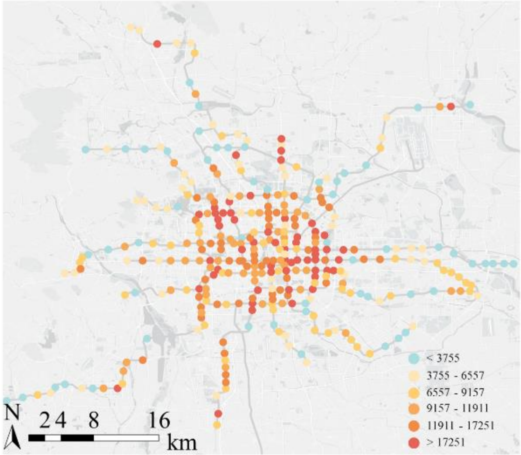

(b)

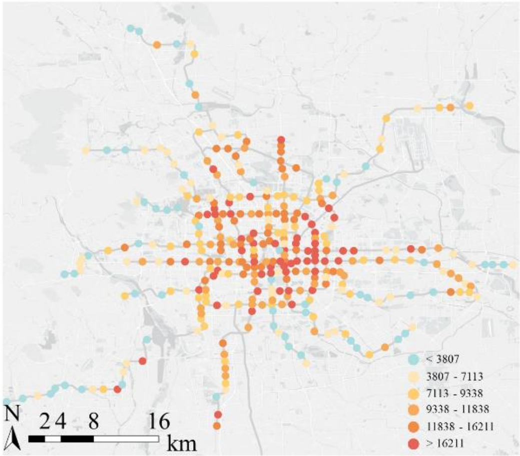

(c)

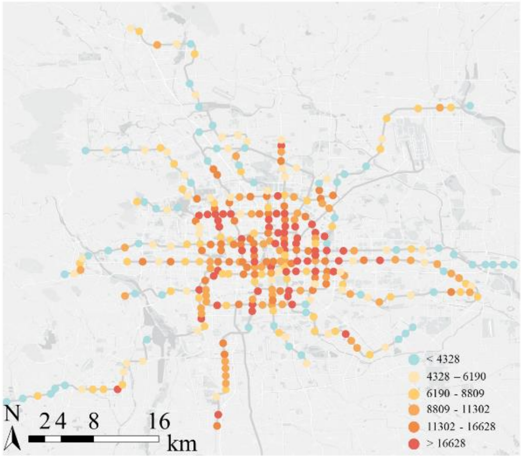

(d)

**Supplementary Figure 4:** Causal estimates under different estimators: (a) S-Learner. (b) T-Learner. (c) X-Learner. (d) Domain Adaptation Learner.

**Supplementary Table 1:** Summary characteristics of overall dataset

.

| Category | Variable | Description | Before the COVID-19 | | During the COVID-19 | |
|---|---|---|---|---|---|---|
| | | | Mean | S.D. | Mean | S.D. |
| Traffic Volume | Ridership | Daily tap-in ridership | 20,094 | 15,982 | 8,383 | 6,744 |
| Demographic | Population | The population of 800 meters around the station | 41,040 | 30,870 | 36,809 | 29,859 |
| Land Use Properties | Bus station | Number of bus station POIs within 800 meters | 9.835 | 5.031 | 9.251 | 4.419 |
| | Accommodation | Number of accommodation POIs within 800 meters | 41.113 | 50.912 | 40.087 | 47.479 |
| | Restaurant | Number of restaurant POIs within 800 meters | 202.339 | 189.692 | 184.525 | 154.115 |
| | Shopping | Number of shopping POIs within 800 meters | 447.508 | 516.613 | 295.358 | 302.834 |
| | Scenery | Number of scenery POIs within 800 meters | 8.639 | 16.228 | 12.681 | 25.771 |
| | Hospital | Number of hospital POIs within 800 meters | 40.942 | 31.862 | 45.442 | 36.526 |
| | Enterprise | Number of enterprise POIs within 800 meters | 291.376 | 365.864 | 167.048 | 178.516 |
| | Residence | Number of residence POIs within 800 meters | 73.511 | 76.769 | 83.063 | 62.324 |
| | House price | Inverse distance-weighted house price within 800 meters (yuan/$m^2$) | 67,715 | 26,343 | 67,771 | 27,845 |
| | Rent | Inverse distance-weighted rent price within 800 meters (yuan/$m^2$/month) | 114.696 | 178.280 | 99.975 | 57.734 |
| Network Metrics | Betweenness | Quantifing the number of shortest paths passing through the station | 0.048 | 0.042 | 0.048 | 0.041 |
| | Closeness | Quantifing how quickly a station can reach other stations | 0.065 | 0.015 | 0.063 | 0.015 |
| | Eigenvector | Assigning importance considering both the number and quality of its neighboring stations | 0.027 | 0.048 | 0.023 | 0.049 |
| | PageRank | An algorithm for calculating the importance of Internet pages | 0.003 | 0.001 | 0.003 | 0.001 |
| Weather | Temperature | Average daily temperature matched by distance (°C) | 26.416 | 2.280 | 26.652 | 2.990 |
| | Relative humidity | Average daily relative humidity matched by distance (%) | 55.746 | 9.753 | 51.765 | 13.442 |
| | Precipitation | Daily cumulative precipitation matched by distance (mm) | 1.067 | 3.461 | 0.549 | 0.915 |
| | Wind speed | Average wind speed matched by distance (m/s) | 1.828 | 0.411 | 2.046 | 0.565 |

Supplementary Table 2: Model comparison

.

| Model | Ordinary Least Squares (OLS) | Geographically Weighted Regression (GWR) | Geographically and Temporally Weighted Regression (GTWR) | LightGBM |
|---|---|---|---|---|
| $R^2$ | 0.475 | 0.723 | 0.732 | 0.941 |

Note: This study calculated the fitting accuracy of four models for learning the data, highlighting the superior robustness conferred by the ensemble learning approach, particularly in comparison to traditional linear models. Accordingly, the LightGBM was selected as main model for this study.

Supplementary Table 3: Relative importance before and during the COVID-19

.

| Feature | Relative importance | |
|---|---|---|
| | Before the COVID-19 | During the COVID-19 |
| Population | 2.78% | 3.67% |
| Bus station | 4.93% | 4.37% |
| Accommodation | 5.45% | 3.32% |
| Restaurant | 29.73% | 24.51% |
| Shopping | 4.98% | 3.77% |
| Scenery | 1.62% | 3.65% |
| Hospital | 1.78% | 4.31% |
| Enterprise | 8.07% | 12.84% |
| Residence | 4.14% | 1.78% |
| House price | 3.30% | 8.79% |
| Rent | 5.83% | 3.99% |
| Betweenness | 13.74% | 3.45% |
| Closeness | 4.48% | 2.70% |
| Eigenvector | 2.84% | 4.54% |
| PageRank | 4.49% | 5.71% |
| Temperature | 0.45% | 4.06% |
| Relative humidity | 0.44% | 2.38% |
| Precipitation | 0.67% | 0.42% |
| Wind speed | 0.28% | 1.77% |
